# A Standard Virus Load Function

**DOI:** 10.1101/2020.06.19.20135814

**Authors:** Thomas Hillen

## Abstract

The idea is to design a simple function that can describe typical virus-load curves without solving a full virus load dynamic model. We present such a standard virus load function and validate it on data from influenza A as well as SARS-CoV-2 infection data for Macaque monkeys and humans. Further, we compare the virus load function to an established *target model* of virus dynamics as presented by A. Smith in [1]. This virus-load function can be used as input to higher level models for the physiological effects of a virus infection, for models for tissue damage, and to develop treatment strategies.

## 1 Introduction

Virus load curves, as reported in [1, 5, 4], have a very typical infection progression (see Figure 1). According to A. Smith [1] the virus infection presents itself in five phases. In the first phase (Phase Ia) the virus quickly infects cells without being detectable. This phase is followed by exponential growth (Phase Ib) until growth shows signs of saturation and a maximum is reached (Phase Ic). A period of slow exponential decline ensues, which we call Phase II. And finally, we often observe a fast decline that leads to virus clearance (Phase III). Depending on the virus and the response of the patient, these phases can be shorter or longer, or are not shown before the patient parishes.

**Figure 1:**
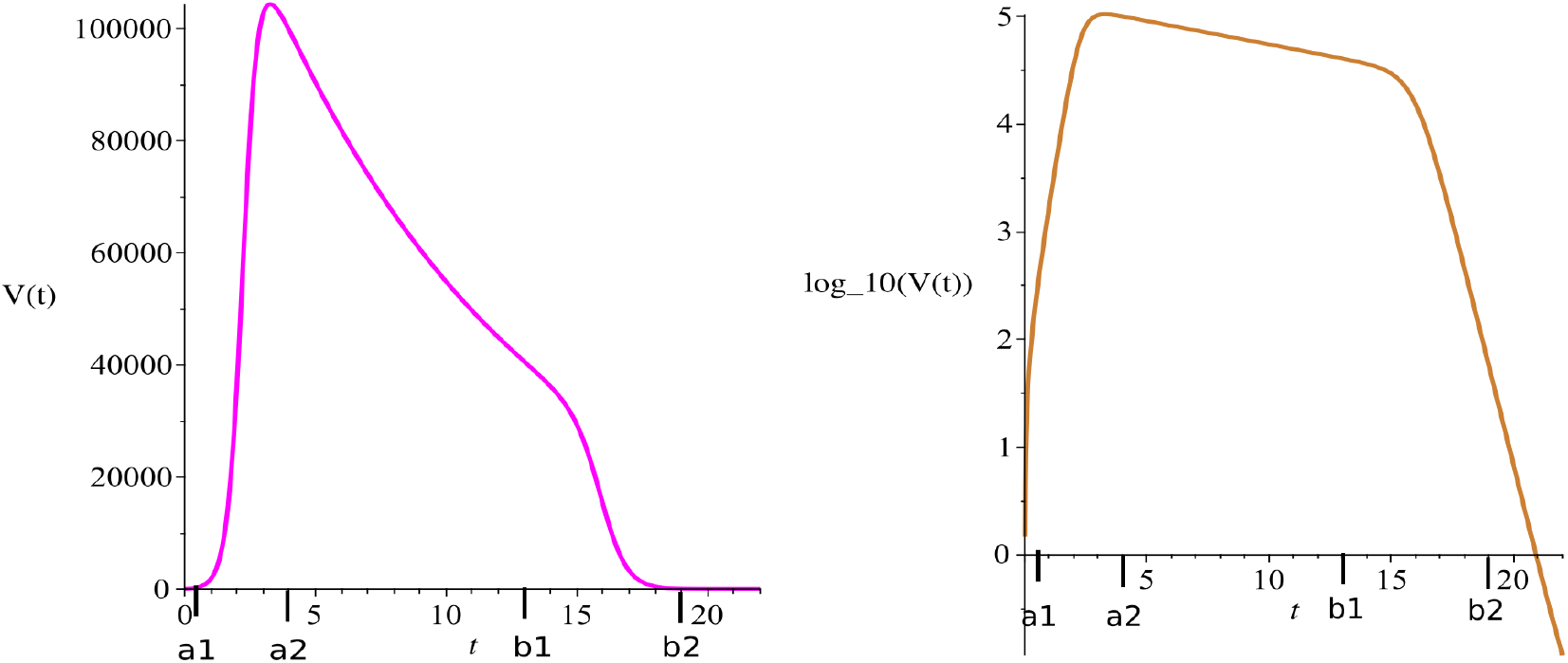
Typical virus load curve. The virus load (“titer”) is usually reported as a dilution value that is needed to infect 50% of a given cell culture, the TCID50 value. Left: absolute scale, right: TCID50-scale (log-scale).

To develop our virus load function (1) we divide the virus load progression into three phases. We consider the Phase I of simgoid increase between time points *a*_1_ and *a*_2_ (see Figure 1). The Phase I includes the three initial phases (Phases Ia, Ib, Ic) of Smith [1] mentioned above. At time *a*_2_ a slow decline of the virus is observed as the immune response kicks in (Phase II between *a*_2_ and *b*_1_ in Figure 1) and finally (Phase III) shows a rather sharp decline once the virus is controlled (between *b*_1_ and *b*_2_). We are guided by the default virus load curve from [8] Figure 2, page 100 and write the virus load curve as a product of three functions, representing the three main phases: 

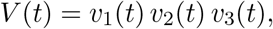

where *v*_1_ describes the initial growth phase between *a*_1_ and *a*_2_, *v*_2_ the intermediate slow decay phase between *a*_2_ and *b*_1_, and *v*_3_ the final decay phase between *b*_1_ and *b*_2_. These are given as sigmoid and exponential functions, respectively 

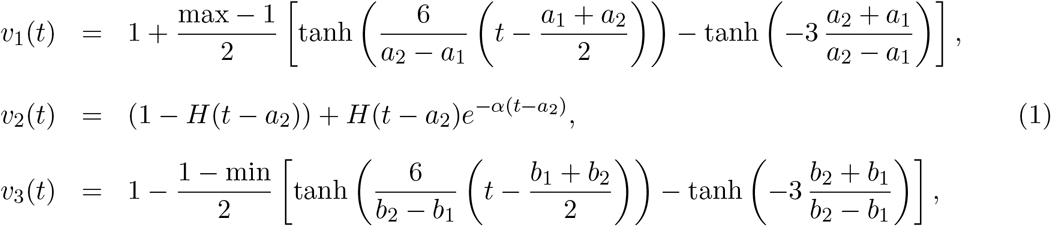

where *H*(*t*) denotes the Heavyside function. The specific form of sigmoid curves for *v*_1_ and *v*_3_ was developed previously by Olobatuyi in [6] in a cancer model. It allows us to define these functions based on intuitive transition threshold values. The value *a*_1_ describes the onset of growth and *a*_2_ a value when saturation is reached, similarly, *b*_1_ denotes the time where decay switches from slow to fast and *b*_2_ is the time when the virus is effectivley eliminated. The parameter *α* describes the intermediate exponential decay rate. In Table 1 we list the values used in Figure 1 and Figure 2 and their meaning.

**Figure 2:**
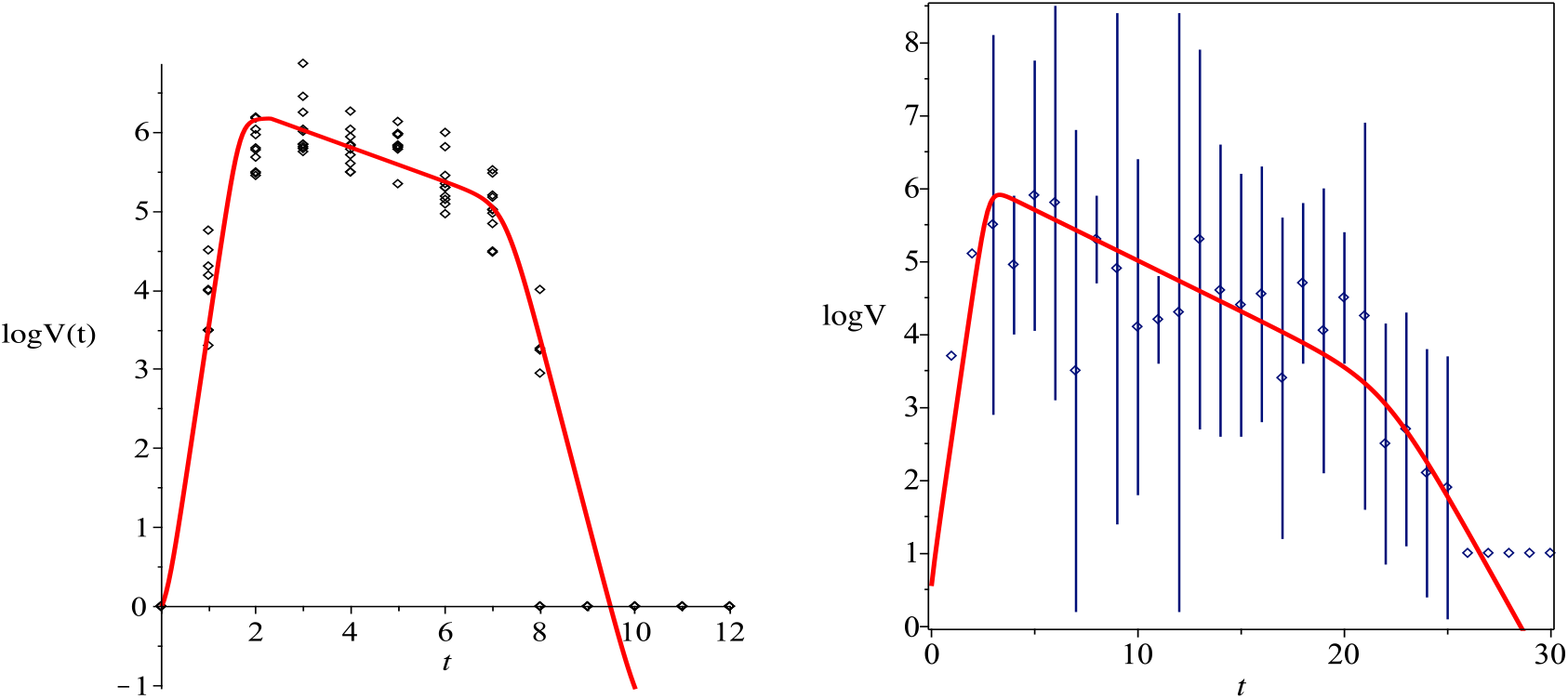
Left: Virus load curve (1) adapted to the data of Influenza A infection in mice. The parameters are listed in Table 1. Right: Human SARS-CoV-2 data from oropharynx saliva samples [3], parameter values are in Table 2.

**Table 1:**
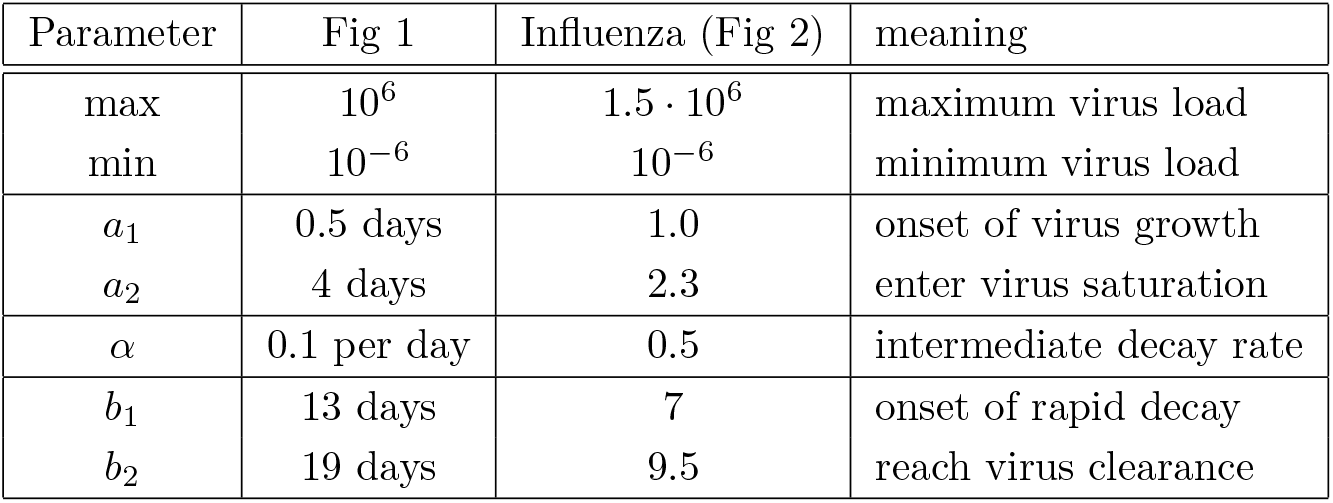
Parameters of the standard virus load function (1). Fig 1. Relates to a typical virus load curve as reported in [1]. Fig 2 parameters are used to fit the influenza data in mice.

| Parameter | Fig 1 | Influenza (Fig 2) | meaning |
| --- | --- | --- | --- |
| max | $10^6$ | $1.5 \cdot 10^6$ | maximum virus load |
| min | $10^{-6}$ | $10^{-6}$ | minimum virus load |
| $a_1$ | 0.5 days | 1.0 | onset of virus growth |
| $a_2$ | 4 days | 2.3 | enter virus saturation |
| $\alpha$ | 0.1 per day | 0.5 | intermediate decay rate |
| $b_1$ | 13 days | 7 | onset of rapid decay |
| $b_2$ | 19 days | 9.5 | reach virus clearance |

**Table 2:** Parameters adapted to the human data (column 2), to the macaque data (columns 3,4,5) and to the target model (column 6).

| Parameter | Human data | Macaque Group 1 | Macaque Group 2 | Macaque Group 3 | Macaque Groups 1,2,3 |
| --- | --- | --- | --- | --- | --- |
| max | 700000 | $6 \cdot 10^6$ | $6 \cdot 10^6$ | $10^6$ | $6 \cdot 10^6$ |
| min | $10^{-6}$ | $10^{-6}$ | $10^{-6}$ | $10^{-6}$ | $10^{-6}$ |
| $a_1$ | 1.5 | 1 | 1 | 1 | 1 |
| $a_2$ | 4 | 2.5 | 2.5 | 2.5 | 2.5 |
| $\alpha$ | 0.43 | 0.52 | 0.55 | 0.43 | 0.5 |
| $b_1$ | 15 | 25 | 25 | 30 | 30 |
| $b_2$ | 29 | 35 | 35 | 35 | 35 |

The hyperbolic tangent function is a sigmoid step function that connects *−*1 to 1. In (1) it has been shifted and scaled such that transitions occur between *a*_1_ and *a*_2_ upwards and between *b*_1_ and *b*_2_ downwards, where the maximum is *max* and the minimum *min*. The ominous value “6” in the argument of the hyperbolic tangent can easily be explained by looking at *a*_1_ = −1 and *a*_2_ = 1. Then 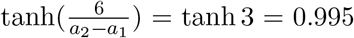 That means that at *a*_2_ a 99.5% of the saturation level is reached. Similar, at *a*_1_ the function is 0.5% above its minimum.

Virus load functions are in high demand in the virus modelling community. For example in [9], a large community of researchers develops an individulal based SARS-CoV-2 physiological model that includes virus infection, virus transmission, immune response, and potential damage to the tissue. The immune response and the tissue complications are directly related to the virus load of the tissue. Our standard virus load function will be a welcome modeling tool to describe

Tissue damage and assess complication risks. Another example of detailed virus modelling is a recent preprint on the impact of SARS-CoV-2 on the renin-angiotensin-system by Pucci et al. [7]. A realistic virus load function is needed as model input in their model. The pharmacological company Pfizer made it their emphasis to develop new treatment strategies and new estimates of side effects, once a Covid-19 treatment becomes available. A virus load function, as presented here, will be a welcome tool to test their ideas.^1^

In the next sections we compare this new virus load function (1) to a very controlled data set of infection of influenza A of mice, and to SARS-CoV-2 data for humans and Macaque monkeys.

## 2 Influenza virus load data

We begin with Influenza A virus load data from [1] as these are the best experimental data available, based on tightly controlled murine experiments. In [1] ten mice were infected with influenza A virus and time series of virus load titers on the log-scale were measured. These data stand out for its tightly controlled set up, allowing to get measurements with minimal spread. In Figure 2 on the left we show those data plus a fit of our virus load curve. The matching can be done by hand, as the parameter values have a clear biological meaning. At this point an optimized fit seems of little value, since many possible curves would fit within the data points and give equally good fits. Here it is more important to get realistic estimates for the model parameters, as they have direct biological meaning about the timing of the infection. The parameter values for the left figure of Figure 2 are listed in Table 1.

## 3 Human SARS-CoV-2

In [3] a cohort of COVID-19 patients from two Hong Kong hospitals was evaluated. 30 patients were screened between Jan. 22, 2020 and Feb 12, 2020, and 23 patients were included in the study. For each patient on a daily basis a multitude of clinical measurements were recorded, including a virus-load measurement. For patients who were not intubated an oropharynx saliva sample was collected. In the early mornings patients were asked to cough up to clear the throat

and the virus load in the saliva was measured. From patients who were intubated a endotrachial aspirate sample was taken. As the ciliary activity of the lung epithelium transports mucus to the posterior oropharyngeal area, these samples give a good indication of the viral activity in the lungs. The data were collected on a daily basis and recorded as mean values and standard deviations. In Figure 2 on the right we show the orophalynx measurements as mean values with error bars. We fit our virus load function (1) to these data as shown on the right of Figure 2 as solid line, and the corresponding model parameters are listed in Table 2. We observe that the initial virus growth phase is rather quick and the virus reaches its carrying capacity within 2.5 days (*a*_2_ *− a*_1_ = 2.5). The virus load reaches a saturation level, which is likely to be related to the innate immune response and it starts a phase of slow decay with a half-life time of 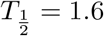 days. After 15 days the virus load drops more quickly, possibly due to the adaptive immune response and at day 29 the virus is cleared.

Again we adapted the curve by hand. The error bars are so wide that a optimization of a mean squared error seems to be futile in this case. We see that the virus load function (1) can conveniently describe all three phases of these dynamics, and, as we will see, the dynamics for macaque monkeys is rather similar.

## 4 Macaque Monkey Data

In [2] nine rhesus macaques monkeys were infected with SARS-CoV-2. Three monkeys (group 1) obtained a high initial virus dose, three monkeys (group 2) a medium initial dose, and three (group 3) a small initial dose. The virus load was measured daily or every other day through a bronchoalveolar lavage probe. The recorded data for each individual group and for all three groups together are shown as dots in Figure 3. All nine monkeys showed only mild disease symptoms and they all fully recovered. Hence the infection cycle here is more indicative of a mild infection, in contrast to the human data considered above.

**Figure 3:**
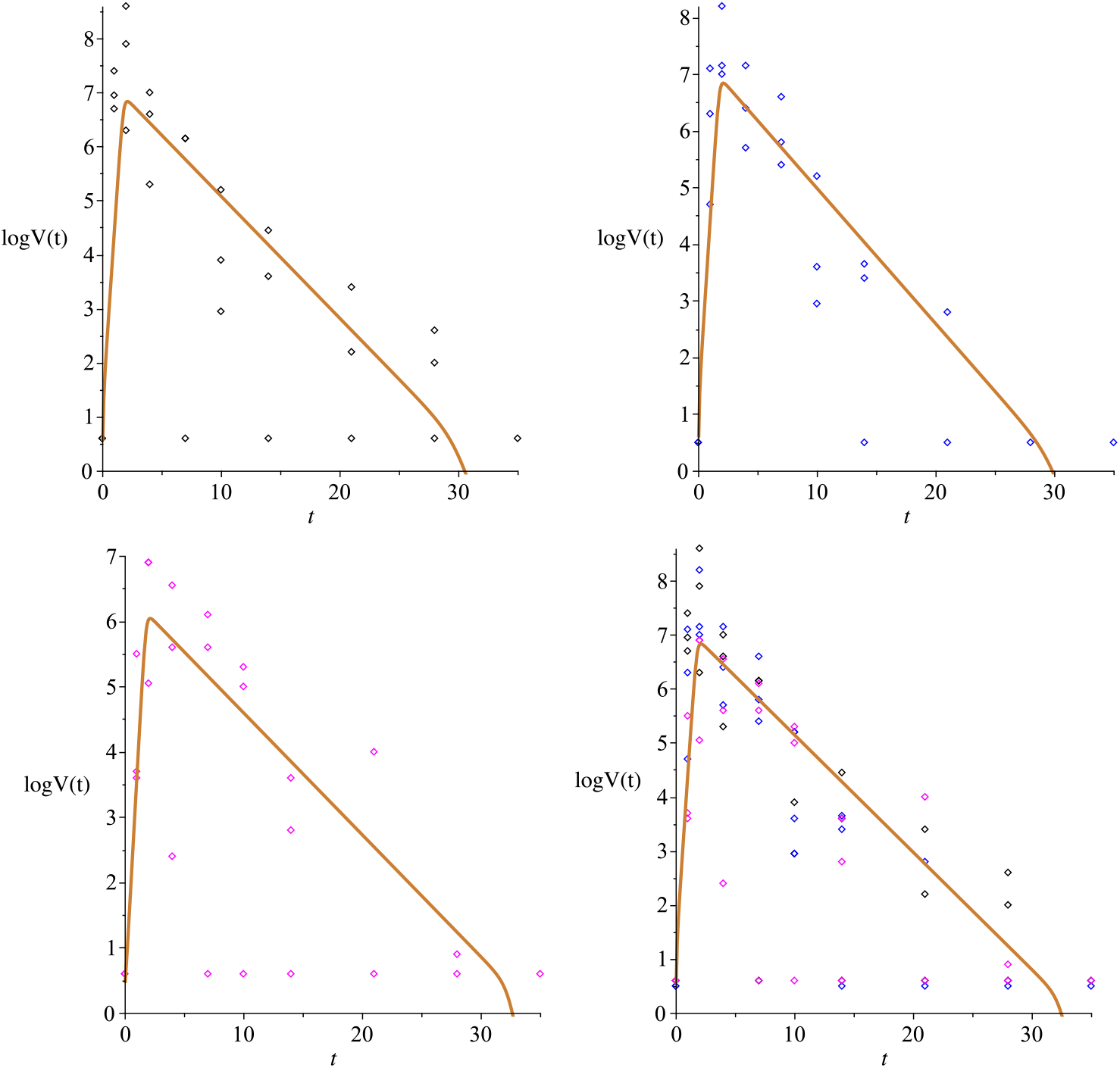
Macaque monkey data: Groups 1,2,3, and all groups together. Data from [2]

In Figure 3 we adapt our virus load function (1) to the rhesus monkey data. We can see in Table 2 that the chosen parameter values show a great level of consistency between the three groups, such that a good fit of all three groups combined is also possible. The characteristic values for the initial virus growth *a*_1_ = 1 and *a*_2_ = 2.5 are identical between the groups, indicating that the amount of initial viral dose is not so important. The time points of the onset of viral decline are in the same order of magnitude *b*_1_ = 25, 25, 30, while the time of virus clearance is *b*_2_ = 35 for all groups. Also the virus decay rates in the intermediate phase of *α* = 0.52, 0.55, 0.43, 0.5 are rather close, corresponding to half-life times of 

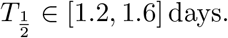

These times include the decay time of the human data of 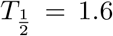 days as reported in the previous section. Hence the intermediate response to the infection appears similar between humans and monkeys. The length of the infection is estimated as 27.5 days for humans and 34 days for the monkeys, where the final decay phase starts significantly earlier in humans *b*_1_ = 15 than in monkeys *b*_1_ = 25 *−* 30. This could be an indication of a higher level adaptive immune response in humans as compared to monkeys.

## 5 Comparison to the Virus-target model of Smith et al

In [1] a virus load model has been fit to the influenza data mentioned above. The so called *target model* comprises of four ordinary differential equations (ODEs) for the target cells *T* (*t*), the infected cells, also called the eclipse phase, *I*_1_(*t*), the infectious cells *I*_2_(*t*) and the virus load *V* (*t*). 

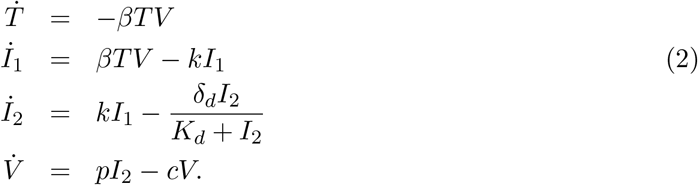

Here *β* is the virus infection rate, *k* the infection maturation rate, *p* the virus production rate, *c* the virus decay rate, *δ*_*d*_ the base decay rate of infectious cells, and *K*_*d*_ the half saturation constant for the decay term of the infectious cells. The parameter values which we use here are listed in Table 3. The parameter values that are published in [1] and their meanings are listed as “Target model 1”. These do not give the best fit to the influenza data from Figure 2 and I am fortunate to have parameter values from a direct communication with the authors of [1], which fit better. We call those alternative choices “Target model 2”.

**Table 3:**
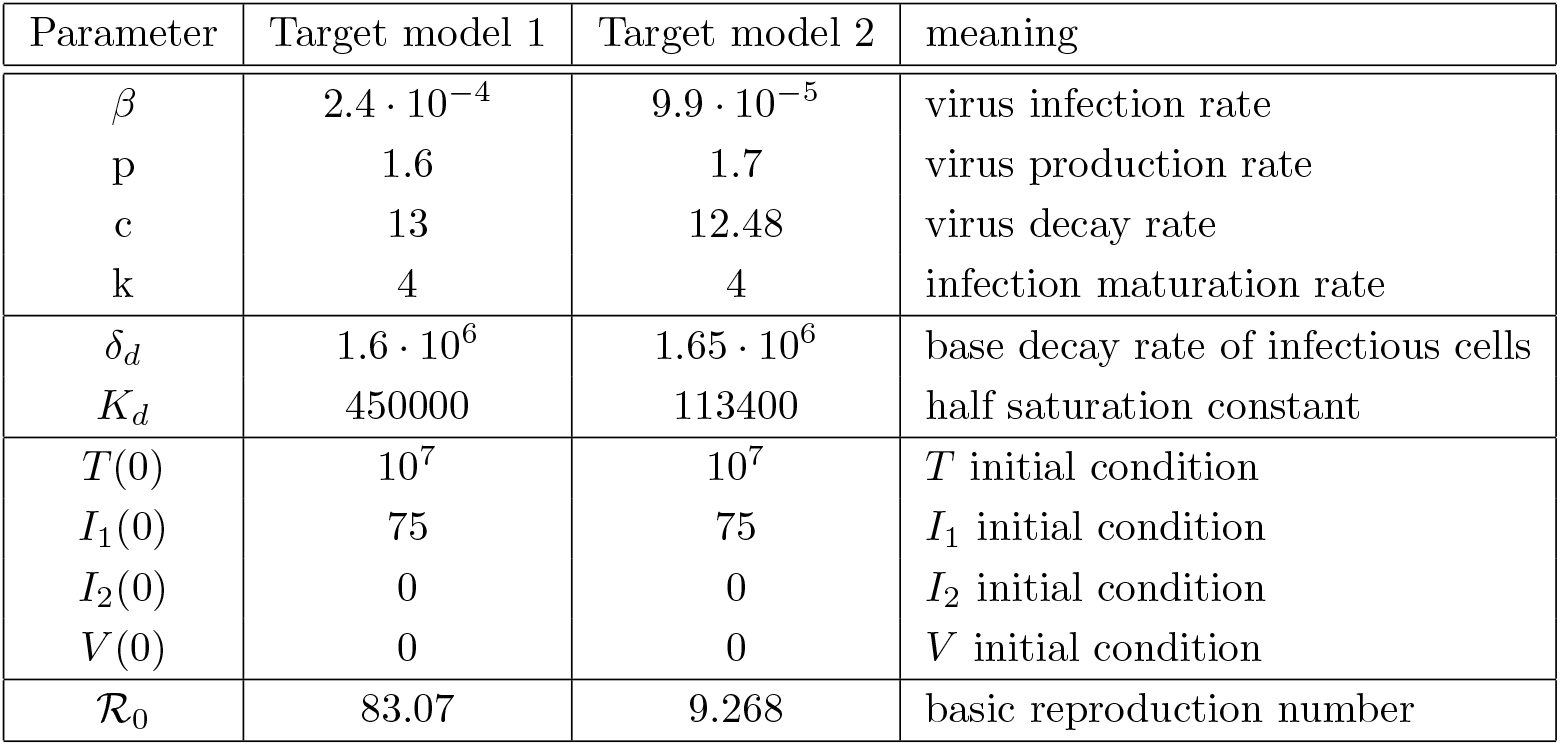
Parameters for the target model ODE (2). Target model 1 refers to the parameter values that are listed in [1]. Target model 2 are those parameter values that produce the best fit to the influenza data from above, and were directly communicated by one of the authors of [1].

| Parameter | Target model 1 | Target model 2 | meaning |
| --- | --- | --- | --- |
| $\beta$ | $2.4 \cdot 10^{-4}$ | $9.9 \cdot 10^{-5}$ | virus infection rate |
| p | 1.6 | 1.7 | virus production rate |
| c | 13 | 12.48 | virus decay rate |
| k | 4 | 4 | infection maturation rate |
| $\delta_d$ | $1.6 \cdot 10^6$ | $1.65 \cdot 10^6$ | base decay rate of infectious cells |
| $K_d$ | 450000 | 113400 | half saturation constant |
| $T(0)$ | $10^7$ | $10^7$ | $T$ initial condition |
| $I_1(0)$ | 75 | 75 | $I_1$ initial condition |
| $I_2(0)$ | 0 | 0 | $I_2$ initial condition |
| $V(0)$ | 0 | 0 | $V$ initial condition |
| $\mathcal{R}_0$ | 83.07 | 9.268 | basic reproduction number |

**Table 4:** Parameters of the virus load function compared to the Influenza data, to the Target model 1 and to the Target model 2.

| Parameter | Influenza data | Target model 1 | Target model 2 |
| --- | --- | --- | --- |
| max | $1.5 \cdot 10^6$ | $1.2 \cdot 10^6$ | $1.3 \cdot 10^6$ |
| min | $10^{-6}$ | $10^{-6}$ | $10^{-6}$ |
| $a_1$ | 1.0 | 0.56 | 0.9 |
| $a_2$ | 2.3 | 1.28 | 2.1 |
| $\alpha$ | 0.5 | 0.28 | 0.4 |
| $b_1$ | 7 | 6.0 | 6.7 |
| $b_2$ | 9.5 | 8.5 | 7.8 |

The basic reproduction number *R*_0_ for this model is given as (see [1]) 

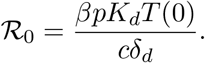

For the two parameter sets that we consider here, we compute *ℛ*_0_ in Table 3. It should be noted that the value of *ℛ*_0_ = 8.8, listed in [1] is not quite accurate.

In Figure 4 we show the virus load curve from the target model as a thick red line and an overlay of the virus load function (1) as a black thin line. Target model 1 is shown on the left and Target model 2 on the right. We see that the virus load function (1) can reproduce both curves with a high level of accuracy. The parameter values are listed in Table 3.

**Figure 4:**
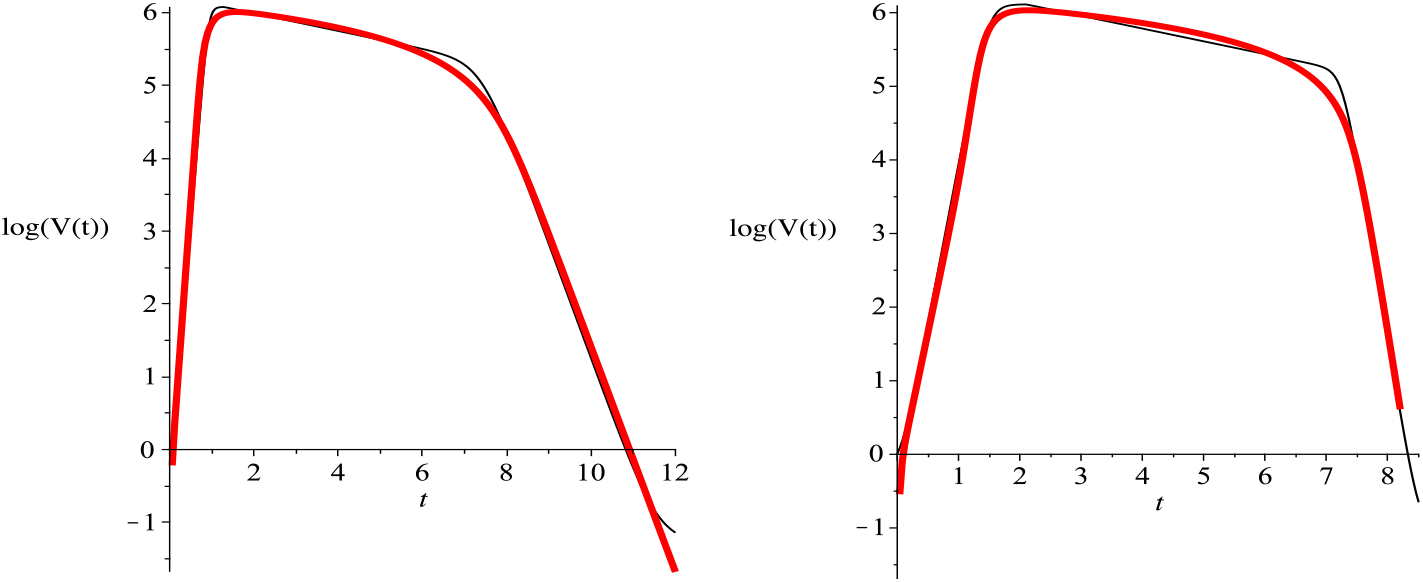
Comparison of the virus load curve (black) to the target model solution (red) for the two cases which we called Target model 1 (left) and Target model 2 (right).

## 6 Conclusion

The explicit form of this new standard virus load function (1) is simple and convenient. The corresponding parameter values all have easily understandable biological meaning, allowing this model to be used quickly and efficiently. We have shown that this virus load function can replicated observed virus load titers from Influenza A in mice and from SARS-CoV-2 in humans and in monkeys. A quick analysis shows already that macaque are good model systems for human infection, where the early and intermediate infection phases are very similar and the final elimination period is quicker in humans.

The virus load function (1) will allow researchers to focus their analysis on higher level effects such as immune responses, tissue damage, ARDS, spread to other organs in the body and for treatment designs.

## Data Availability

All data have been previously published. Full references are given.

## Acknowledgements

I am grateful to J. Newby and A. Smith for helpful comments to an early version of this idea. I am also most grateful to A. Smith to freely sharing the influenza virus load data, which her group collected in painstakingly tedious work.

## Footnotes

1 Pfizer has, due to proprietary reasons, not yet published on Covid-19 modelling. My statements above are based on personal communications.

## References

[1] V. Bernauerova A.M. Smith A.P. Smith, D.J. Moquin. Influenza virus infection model with density dependence supports biphasic viral decay. Frontiers in Microbiology, 9:1–10, 2018.

[2] A. Chandrashekar, J. Liu, A.J. Martinot, and etal. Sars-cov-2 infection protects against rechallenge in rhesus macaques. Science, page DOI: 10.1126/science.abc4776, 2020.

[3] WS. Leung et al. K. Kai-Wang, O Tak-Yon Tsang. Temporal profiles of viral load in posterior oropharyngeal salive samples and serum antibody responses during infection by sars-c0v-2: an observational cohort study. Lancet, 20:568–574, 2020.

[4] S.D. Solomon O. Vardeny M. Madjid, P. Safavi-Naeini. Potential effects of coronaviruses on the cardiovascular system. JAMA Cardiology, DOI:10.1001/jamacardio.2020.1286, 2020.

[5] L.C. Lane D.J. Moquin P. Vogel S. Woolard A. M. Smith M.A. Myers, A.P. Smith. Dynamically linking influenza virus infection with lung injury to predicy disease severity. bioRxiv, DOI:10.1101/555276, 2019.

[6] O. Olobatuyi, G. de Vries, and T. Hillen. A reaction-diffusion model for radiation-induced bystander effects. J. Math. Biol., 75, 2017.

[7] F. Pucci, P. Bogaerts, and M. Rooman. Modeling the molecular impact of the sars-cov-2 infection on the renin-angiotensin system. 2006.02772v1, 2020.

[8] Amber M. Smith. Host-pathogen kinetics during influenza infection and coinfection: insights from predictive modeling. Immunological Reviews, 285:97–112, 2018.

[9] Y. Wang, G. An, A. Becker, C. Cockrell, N. Collier, M. Craig, CL. Davis, J. Faeder, AN. Ford Versypt, JF. Gianlupi, JA. Glazier, R. Heiland, T. Hillen, MA. Islam, A. Jenner, B. Liu, PA. Morel1, A. Narayanan, J. Ozik, P. Rangamani, JE. Shoemaker, AM. Smith, and P. Macklin. Rapid community-driven development of a sars-cov-2 tissue simulator. bioRxiv, 2020.04.02.019075, 2020.

